# Research on the Application of AI Agent Technology in Quality Defect Root Cause Analysis of Central Sterile Supply Department

**DOI:** 10.64898/2026.04.29.26351275

**Authors:** Min Yi, Xinghua Zhang, Dan Zhao, Qing Zhao

## Abstract

**Objective:** To explore the application effect of AI agent-assisted root cause analysis in the management of quality inspection defects in the Central Sterile Supply Department (CSSD) and to systematically compare it with traditional manual analysis methods.

**Methods:** A retrospective case simulation comparative study was conducted. Thirty typical CSSD quality inspection defect cases were selected. Root cause analysis was performed independently by an AI agent-assisted analysis group and a traditional manual analysis group. Using the consensus results of a high-level expert panel as the “gold standard,” a quantitative comparison was made across four dimensions: analysis quality, efficiency, practicality, and process experience, employing t-tests and Mann-Whitney U tests.

**Results:** Compared with the traditional method, the AI-assisted group demonstrated a significantly higher root cause identification accuracy rate (85.6% vs. 72.3%, P<0.001), superior analysis depth (4.4 points vs. 3.6 points, P<0.001), significantly shorter time consumption per case analysis (18.5 minutes vs. 35.2 minutes, P<0.001), and generated more innovative corrective measures (1.8 items/case vs. 0.7 items/case, P<0.001). There was no statistically significant difference between the two groups regarding the feasibility of the proposed measures (4.0 points vs. 4.2 points, P>0.05).

**Conclusion:** The AI agent-assisted root cause analysis method significantly improves the accuracy, depth, and efficiency of analyzing quality inspection defects in the CSSD and facilitates the discovery of more innovative solutions, demonstrating high application value and promotion potential.

**Implications for Nursing Management:** This study provides empirical evidence that AI agent technology can be integrated into CSSD quality management to enhance defect analysis efficiency and accuracy. Nursing managers should consider adopting AI-assisted tools to standardize root cause analysis processes, reduce reliance on senior staff experience, and enable faster, data-driven decision-making. The reduced training burden and improved novice performance suggest that AI can help address workforce skill gaps. Future implementation should focus on human-AI collaboration, with managers ensuring adequate training, maintaining human oversight, and periodically updating the knowledge base to reflect local clinical contexts.

## Introduction

In recent years, with the rapid development and widespread application of large-scale AI agent technology, the practice of AI Agents in medical quality control has gradually attracted significant attention from both industry insiders and outsiders. In foreign research practices, artificial intelligence technology has mostly focused on utilizing intelligent algorithms to build predictive models for infection control ^[1]^. However, the application of AI Agents in assisting quality control personnel in root cause analysis of defects in the sterile supply chain remains in its preliminary exploratory stage. In contrast, domestic research and applications have placed greater emphasis on the infrastructure construction of information systems ^[2]^, yet have not fully achieved dynamic root cause inference capabilities for quality issues. Currently, mainstream root cause analysis methods predominantly rely on traditional techniques such as fishbone diagrams, 5 Why analysis, and brainstorming. These methods heavily depend on subjective judgments based on human experience, resulting in delayed response times, difficulty in achieving early risk warnings, and often insufficient analysis depth due to cognitive limitations of personnel ^[3]^. To address these issues, particularly the pain points in the root cause analysis process of Central Sterile Supply Departments (CSSD) such as prolonged cycles, the need to involve senior staff, high labor costs, and incomplete improvement suggestions, this paper proposes an innovative “AI + Quality Inspection” approach by introducing AI Agent technology to construct intelligent analytical models with dynamic reasoning capabilities. This model conducts in-depth mining and fusion processing of historical and real-time operational data, systematically analyzing potential influencing factors across six dimensions: personnel, machinery, materials, methods, environment, and management. By integrating knowledge graphs and Retrieval Augmented Generation (RAG) technology, it achieves real-time, automated root cause localization for defect records. The system instantly generates actionable improvement recommendations and automatically matches similar scenarios from a historical case library to predict potential risks, recommending corresponding preventive strategies. This approach enhances the efficiency and accuracy of root cause analysis, driving the transformation of medical quality control from experience-driven to data intelligence-driven, effectively reducing defect response time to minutes. Through continuous learning and dynamic optimization of multi-source heterogeneous data by AI agents, the system not only reduces reliance on expert experience but also significantly improves the consistency and traceability of analytical results ^[4]^. In practical operations, the system automatically identifies typical defect patterns such as instrument cleaning residues, poor packaging seals, and sterilization parameter deviations, precisely locating them to specific processes and responsible nodes ^[5]^. Combined with real-time early warning mechanisms and closed-loop improvement feedback, it enables full lifecycle management of quality issues. Future applications may extend to cross-institutional collaborative analysis, contributing to the establishment of a standardized, intelligent paradigm for medical quality control.

## 1 Materials & Methods

### 1.1 Materials Source

The data for this study were derived from the quality management records of a tertiary Grade A hospital’s sterilization supply center from January 2023 to December 2025. Through systematic review, a preliminary screening identified 50 reported quality control defect incidents. The inclusion criteria required complete documentation of the incident process, including key information such as incident entity details, process traceability data, and environmental/contextual factors. Cases with severe information gaps or extremely rare isolated incidents were excluded. Each case underwent standardized de-identification processing to form a case information package encompassing dimensions such as incident description, process data, environmental factors, and personnel information ^[2]^. Ultimately, 30 representative defect cases were selected to construct the cost study case library. The defect types covered four major categories: substandard instrument cleaning (6 cases), packaging defects (18 cases), non-compliant sterilization parameters (2 cases), and instrument functional damage (4 cases), ensuring representativeness and diversity of the cases.

### 1.2 Method

#### 1.2.1 Dataset

Both groups analyzed the same batch of cases, documenting the complete analytical process, including proposed potential causes, final root cause conclusions, and recommended measures. Both groups were provided with identical case information and the same analytical timeframe, with each case not exceeding 15 minutes. The core collected data included: 1) Event ontology information: specific defect description, discovery stage, and severity classification; 2) Process traceability data: involved operators, information on surgical instruments or instrument packages processed, device numbers and status, consumable batches, and sterilization or cleaning procedure parameters at the time; 3) Environmental and contextual information: shift occurrence, workload, and recent records of related training or protocol changes. All information was consolidated into a uniformly designed case information collection form and independently verified by two researchers to ensure accuracy. The standardized case packages formed were used as common input materials for subsequent traditional analysis and AI-assisted analysis, ensuring baseline consistency in the comparative experiment. This study protocol has been reviewed and approved by the hospital ethics committee, and all data usage adhered to the principles of anonymization and confidentiality.

#### 1.2.2 Specific Methods

##### 1.2.2.1 Experimental Group (Agent Group)

An AI agent for root cause analysis of quality control in disinfection supply centers was developed based on Doubao. A quality control personnel proficient in AI operations entered case information to guide the AI in completing the analysis.

A specialized AI agent for root cause analysis of quality control in Central Sterile Supply Department (CSSD) was constructed based on the “Doubao” large language model platform. This agent aims to simulate and enhance the systematic thinking and reasoning capabilities of senior quality analysis experts, with its core being the development of an intelligent analytical model equipped with dynamic perception, autonomous reasoning, and iterative validation capabilities ^[5]^.

**(1) Agent Architecture and Core Components:** This intelligent agent adopts a modular architecture, primarily consisting of three layers: the Perception and Input Layer, the Analysis and Inference Engine Core Layer, and the Output and Validation Layer. **The Perception and Input Layer** is operated by a quality control personnel familiar with AI operations, who inputs case information. This personnel structures standardized case information (including event descriptions, process data, environmental parameters, etc.) into the system and, based on preliminary analysis results, follows preset prompt words and thought chain instructions to guide the AI in further inquiring about key details (e.g., “Please further analyze the defect frequency of this medical device package over the past three months.”). **The Analysis and Inference Engine Core Layer** integrates multiple technologies such as multidimensional factor correlation analysis, knowledge graph technology, retrieval enhancement (RAG), and thought chain reasoning (CoT) to achieve deep analysis. The multidimensional factor correlation analysis module is based on a six-dimensional framework of “people, machines, materials, methods, environment, and management,” establishing a structured root cause analysis workflow within the agent. It automatically categorizes input information into corresponding dimensions and dynamically constructs association networks between factors to avoid common dimension omissions in traditional analysis. Knowledge graph technology employs embedded vectorization techniques to process industry standards such as CSSD professional knowledge, hospital disinfection supply center management specifications ^[10]^, cleaning, disinfection, and sterilization operation standards ^[11]^, and cleaning, disinfection, and sterilization effect monitoring standards ^[12]^, as well as related equipment principles, standard operating procedures, material characteristics, and microbiological knowledge. It also vectorizes and stores knowledge graph vector data and historical defect case databases. This study employs Retrieval Augmentation (RAG) technology during reasoning processes, where the intelligent agent dynamically retrieves analogous scenarios and root causes from a case library to provide empirical references for current analyses. Simultaneously, it retrieves relevant principles and standards from the knowledge graph to ensure the professionalism and compliance of analytical recommendations. The Coherence of Thought (CoT) technology leverages the logical reasoning capabilities of large language models. The intelligent agent integrates digital expert knowledge to trace and evaluate the relevance network, automatically generating a reasoning chain from surface phenomena to root causes while proposing multiple candidate root cause hypotheses. **The Output and Validation Layer** outputs structured reports through the intelligent agent, including: the most probable root cause (if multiple causes exist, ranked by confidence level), a concise logical chain demonstrating the reasoning process, improvement suggestions based on the knowledge graph and historical cases (e.g., “Adjust the cleaning and packaging procedures for this model of laparoscopic instrument kits, referencing similar cases from X month 2025”), and identified potential similar risk points.

(2) Operational workflow analysis : During the experiment, a quality control specialist inputs case-specific data into the AI root cause analysis system of the Central Sterile Supply Department. The system first performs preliminary analysis and categorization. With human guidance, users input specific keywords or select inquiry directions to initiate multi-round analytical cycles. Through iterative refinement, the system progressively identifies critical contradictions until reaching a stable conclusion. The entire process requires documentation of key AI inquiry points, referenced case studies or knowledge entries, and the step-by-step formation of final conclusions.

##### 1.2.2.2 Control group (traditional RCA method group)

Composed of 3-5 CSSD quality control personnel, head nurses, or hospital infection specialists with over 5 years of experience, using traditional tools such as fishbone diagrams and the 5Why method for analysis ^[9]^.

##### 1.2.2.3 Establishment of Evaluation Criteria

A high-level expert panel (comprising senior CSSD managers, infection control authorities, and quality management experts, with no fewer than 5 members) shall conduct blind analysis of each case to generate the final “Root Cause Reference List” and “Improvement Measure Plan” as the evaluation benchmark.

##### 1.2.2.4 Outcome Evaluation

Submit the output results of both groups to the expert panel (with group information concealed) for quantitative scoring and qualitative assessment based on predefined observation indicators.

#### 1.2.3 Observation Indicators

(1) Analysis quality indicators include four aspects: root cause identification accuracy rate, root cause analysis depth, analysis comprehensiveness, and logical chain integrity. The root cause identification accuracy rate is calculated by comparing the analysis results with the root causes identified by the expert panel, using the formula (overlapping root causes / expert panel-identified root causes) × 100%. Root cause analysis depth is scored based on the number of identified cause levels and whether it touches management systems or process design levels, with a 5-level scoring standard: 1-surface-level analysis, 2-intermediate-level analysis, 3-deep-level analysis, 4-systematic analysis, 5-fundamental institutional improvement. Analysis comprehensiveness evaluates whether it covers multidimensional factors such as people, machines, materials, methods, and environment, assessing the breadth of identified root causes, particularly their ability to capture multi-factor coupling issues, using a 0-10 point scale. Logical chain integrity evaluates the coherence and consistency of the reasoning process, assessing whether there are breaks or jumps in the logical chain, rated on a 1-5 point scale. Due to the non-normal distribution characteristics of root cause analysis depth, non-parametric testing methods are used for statistical analysis. The Mann-Whitney U test is employed for inter-group comparisons, with its Z-value used to measure the significance of differences in analysis depth between groups, where P<0.05 indicates statistically significant differences. For the remaining indicators, the root cause identification accuracy, analytical comprehensiveness, and logical chain integrity were analyzed using independent samples t-tests. Results were expressed as mean ± standard deviation, with the t-value indicating the significance level of differences between groups, where P<0.05 was considered statistically significant. Inter-group comparisons were performed using SPSS 25.0 software. The detailed analysis methods are shown in Table 1.

**Table 1.** Analysis of quality indicator description.

| quota | detailed description | measuring method |
| --- | --- | --- |
| root cause identification accuracy | The degree of overlap between the root causes identified and the expert panel's evaluation criteria. | Calculate (repeated root factor / root factor determined by the expert panel) × 100% |
| root cause analysis depth | Whether it can be traced back to systemic causes (such as processes, policies, training) rather than remaining at the level of individual errors. | The expert panel conducted a 5-level scoring system to evaluate the depth of the analytical pathway. |
| analytic comprehensiveness | Whether it covers all potential dimensions such as people, machine, material, method, environment, measurement, etc. | Compare the number of dimensions in the statistical analysis report with the expert panel's coverage |
| logical chain integrity | The chain of reasoning from the phenomenon to the root cause is clear, continuous and without logical break. | The expert panel conducted a 5-level scoring system to evaluate the rigor of the logical chain. |

(2) The analysis efficiency metrics encompass average single-case analysis duration and root cause identification rate per unit time. The former measures the time span from task initiation to submission of a complete analysis report, recorded in minutes. The latter quantifies valid root cause identification within 20-minute intervals, serving as an indicator of cognitive density and productivity. Both metrics are analyzed by comparing the averages of two teams, with quality metrics integrated to evaluate the comparative advantages of different analytical approaches. Detailed methodologies are presented in Table 2.

**Table 2.** Analysis of efficiency indicators.

| quota | detailed description | measuring method |
| --- | --- | --- |
| Average time per case analysis | Time from receiving a case to generating a complete report. | Timed recording, comparing the means of both groups |
| root cause count per unit time | The number of valid possible causes submitted within the specified time (e.g., the first 20 minutes). | Number of valid possible causes per time |

(3) The implementation feasibility indicators of the proposal include the feasibility of improvement measures and the innovation of measures. The feasibility of improvement measures primarily evaluates the possibility of implementing the proposed scheme under existing resources and conditions, including the rational matching of factors such as cost, manpower, and time. The innovation of measures examines whether the proposal breaks through traditional thinking and proposes new methods with foresight and promotion value. The scoring criteria for the feasibility of improvement measures are as follows: Level 1: Extremely unrealistic, Level 2: Relatively unrealistic, Level 3: Basically feasible, Level 4: Relatively easy to implement, Level 5: Completely feasible. The scoring criteria for the innovation of measures are Level 5: Level 1—Adopting conventional practices without innovation; Level 2—Slightly adjusting conventional methods; Level 3—Proposing partial improvement ideas based on actual conditions; Level 4—Introducing new perspectives or cross-domain methods with certain foresight; Level 5—Creatively proposing new models or paths that are significantly superior to existing practices. A comprehensive score is also calculated based on the number of unique and valuable measures identified by expert judgment. The scoring process is independently completed by three senior experts, and the final average is taken to reduce subjective bias. All indicator data are entered into the system through a double-blind method to ensure objective and fair evaluation. Data analysis is performed using SPSS 25.0 for descriptive statistics and inter-group difference testing, with P<0.05 considered statistically significant. Detailed analysis methods are shown in Table 3.

**Table 3.** Indication of scheme feasibility.

| quota | detailed description | measuring method |
| --- | --- | --- |
| Feasibility of improvement measures | The feasibility and cost-effectiveness of the proposed measures in clinical practice were evaluated. | The evaluation is conducted by frontline CSSD managers on a 5-level scale: Level 1 (extremely unrealistic or non-innovative), Level 2 (relatively unrealistic or low innovation), Level 3 (basically feasible with some innovation), Level 4 (relatively easy to implement with good innovation), and Level 5 (fully feasible with significant innovation value). |
| Innovative measures | Whether it has put forward the novel and systematic solution which is easy to be ignored in the traditional analysis. | The number of "unique and valuable measures" identified by the expert panel. |

The process and user experience metrics encompass three key aspects: process standardization, cognitive load on users, learning curve, and usability. Detailed analysis methods are presented in Table 4. Process standardization evaluates whether the analysis follows the predefined root cause analysis workflow, including step completeness and tool usage correctness. Review experts conduct a 5-level scoring system based on process execution records: 1 indicates severe deviation from the process, 2 partial step omission or improper tool use, 3 basic adherence with minor flaws, 4 satisfactory execution with appropriate tool application, and 5 fully standardized execution demonstrating process optimization. Cognitive load is assessed using the NASA-TLX (NASA Task Load Index, a subjective workload evaluation tool developed by NASA that measures psychological and physiological stress through multidimensional questionnaires) to gauge operators ‘mental pressure and task difficulty during analysis. Learning curve and usability are evaluated by recording new users’ first-time independent analysis time and error rates, combined with System Usability Scale (SUS) scores for comprehensive assessment ^[14]^.

**Table 4.**
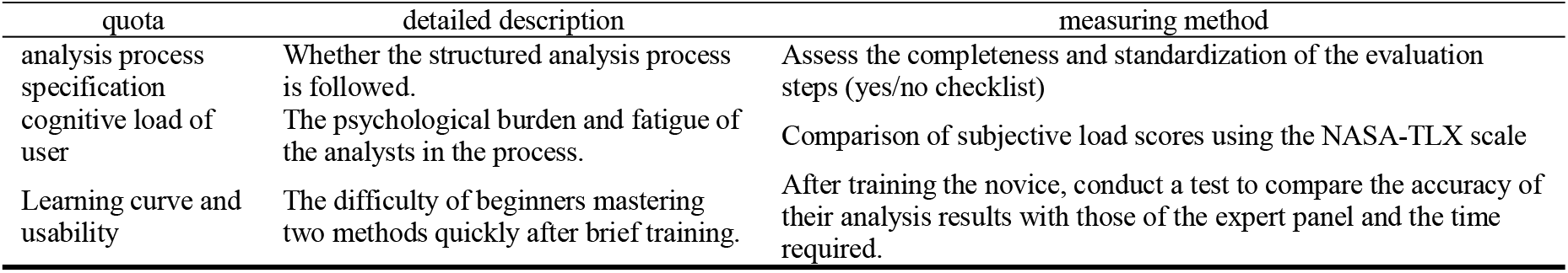
Process and User Experience Metrics.

| quota | detailed description | measuring method |
| --- | --- | --- |
| analysis process specification | Whether the structured analysis process is followed. | Assess the completeness and standardization of the evaluation steps (yes/no checklist) |
| cognitive load of user | The psychological burden and fatigue of the analysts in the process. | Comparison of subjective load scores using the NASA-TLX scale |
| Learning curve and usability | The difficulty of beginners mastering two methods quickly after brief training. | After training the novice, conduct a test to compare the accuracy of their analysis results with those of the expert panel and the time required. |

### 1.3 Statistical Analysis

After the experiment, statistical methods were employed to compare the differences between the two groups. For quantitative data such as accuracy and time consumption, if the data followed a normal distribution, the paired t-test was used to compare the differences between the groups; if not, the sign-rank test was applied. For scoring data, the Mann-Whitney U test was utilized. The effect size was calculated to assess the practical significance of the differences, rather than merely their statistical significance. Data analysis was performed using SPSSAU software, with a P-value <0.05 considered statistically significant.

### 1.4 Ethical Consideration

This study was approved by the Ethics Committee of Sichuan Provincial People’s Hospital(伦 审 (研)2026 年 第 383 号). All data were anonymized and de-identified prior to analysis. The requirement for informed consent was waived by the ethics committee because the study involved retrospective analysis of quality management records with no patient-identifiable information. The research was conducted in accordance with the Declaration of Helsinki.

## 2 Results

### 2.1 Comparison of quality indicators between the two groups

Table 5 demonstrates that the AI-assisted group significantly outperformed the traditional method group in root cause identification accuracy, analytical depth, and comprehensiveness (p<0.001), with superior logical chain integrity (p=0.045). The root cause analysis Z-score of 3.89 further confirms that AI agent assistance markedly enhances systematic rigor, particularly excelling in deep attribution of complex issues. This indicates that AI agents not only improve analytical efficiency but also expand human cognitive boundaries in both breadth and depth of thought.

**Table 5.**
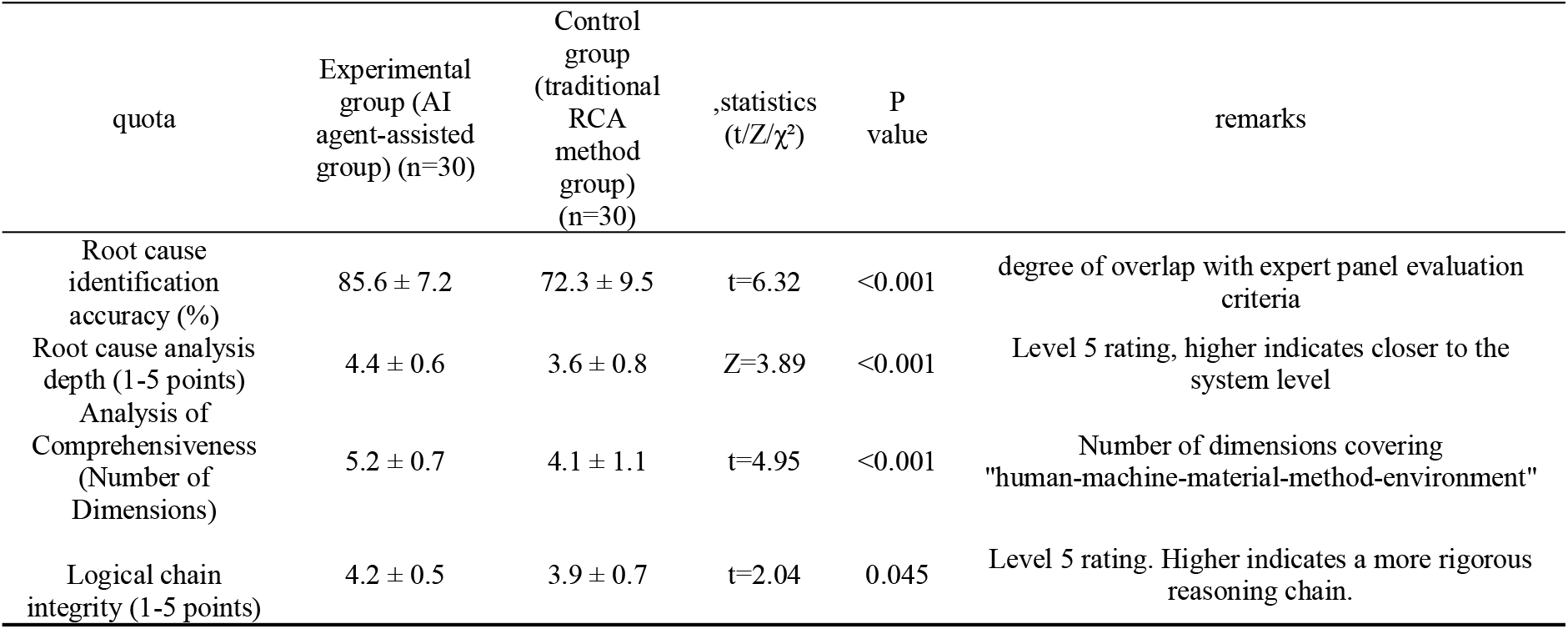
Analyze the comparison results of quality indicators.

### 2.2 Comparison of efficiency indicators between the two groups

Table 6 demonstrates that the AI-assisted group significantly outperforms the traditional method group in three key metrics for analysis efficiency: average single-case analysis time, root cause identification rate per unit time, and time to first core root cause identification (p<0.001). The Cohen’s d effect sizes for these metrics are 2.14,1.56, and 1.98 respectively, indicating substantial efficiency gains from AI assistance. Particularly in complex cases, AI rapidly pinpoints critical clues, shortens problem localization time, and enhances effective hypothesis generation per unit time, providing robust support for real-time decision-making.

**Table 6.** Compare analysis efficiency metrics.

| quota | Experimental group (AI agent-assisted group) (n=30) | Control group (traditional RCA method group) (n=30) | statistics (t/Z/$\chi^2$) | P value | remarks |
| --- | --- | --- | --- | --- | --- |
| Average time per case analysis (minutes) | 18.5 ± 4.2 | 35.2 ± 8.7 | t=-10.24 | <0.001 | Total time from case receipt to report completion |
| Root cause count per unit time (per 20 minutes) | 9.8 ± 2.1 | 6.3 ± 1.8 | t=7.12 | <0.001 | Number of valid possible causes proposed within the first 20 minutes |
| First touch core root cause time (minutes) | 8.3 ± 3.1 | 19.6 ± 6.5 | t=-9.35 | <0.001 | Time from the start of analysis to the first identification of critical root causes |

### 2.3 Comparison of Practicality Indicators Between the Two Treatment Groups

Table 7 demonstrates that the AI-assisted group significantly outperformed the traditional method group in both the innovativeness and systematicness of proposed solutions (p<0.001 and p=0.001), particularly in generating unique and systematically improved proposals. This indicates that AI not only enhances analytical efficiency but also elevates the quality and depth of solutions. No significant difference was observed between the two groups in terms of the feasibility of improvement measures (p=0.141), suggesting that AI-assisted methods did not compromise the practicality of solutions. Notably, by integrating multi-source data and historical cases, AI can identify systemic flaws often overlooked by traditional methods, thereby proposing more forward-looking optimization paths. This capability is particularly critical in complex production environments, as it reduces reliance on human experience while improving decision consistency and scientific rigor.

**Table 7.**
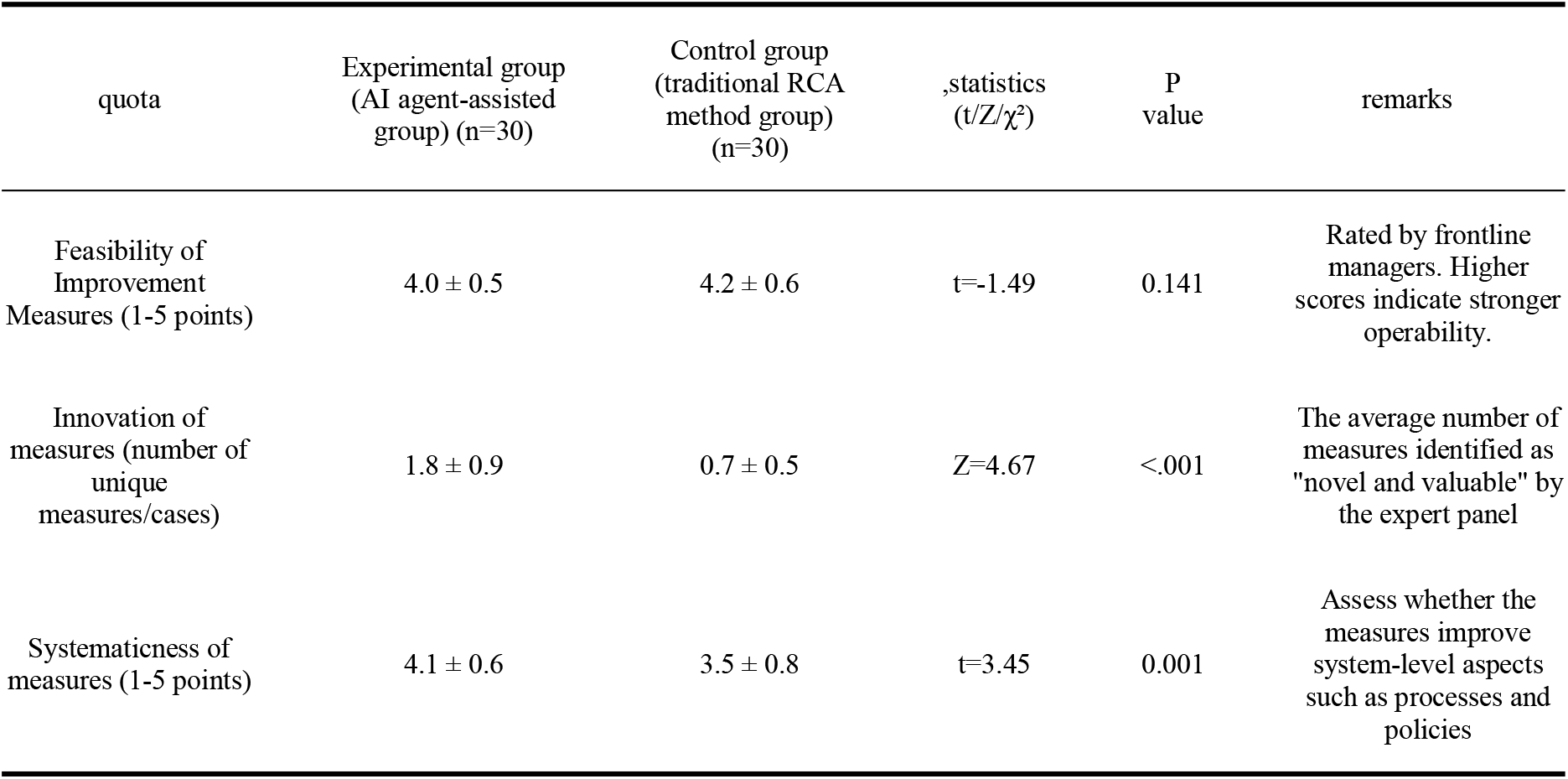
Comparison results of the plan’s practicality indicators.

| quota | Experimental group<br>(AI agent-assisted<br>group) (n=30) | Control group<br>(traditional RCA<br>method group)<br>(n=30) | statistics<br>(t/Z/$\chi^2$) | P<br>value | remarks |
| --- | --- | --- | --- | --- | --- |
| Feasibility of<br>Improvement<br>Measures (1-5 points) | 4.0 ± 0.5 | 4.2 ± 0.6 | t=-1.49 | 0.141 | Rated by frontline<br>managers. Higher<br>scores indicate stronger<br>operability. |
| Innovation of<br>measures (number of<br>unique<br>measures/cases) | 1.8 ± 0.9 | 0.7 ± 0.5 | Z=4.67 | <.001 | The average number of<br>measures identified as<br>"novel and valuable" by<br>the expert panel |
| Systematicness of<br>measures (1-5 points) | 4.1 ± 0.6 | 3.5 ± 0.8 | t=3.45 | 0.001 | Assess whether the<br>measures improve<br>system-level aspects<br>such as processes and<br>policies |

### 2.4 Comparison of Process and User Experience Indicators Between Two Groups

Table 8 demonstrates that the AI-assisted group significantly outperformed the traditional method group in process standardization, cognitive load, result proximity, and user satisfaction (p<0.05), with effect sizes ranging from 0.93 to 2.17. This indicates that AI not only enhances process standardization but also substantially reduces users ‘psychological burden, particularly facilitating new users to rapidly achieve expert-level analytical proficiency ^[6]^. The tool’s intuitive design and guidance improve operational confidence and user experience, revealing that AI-assisted systems optimize analytical pathways while redefining cognitive efficiency and subjective value perception in human-computer collaboration, thereby demonstrating dual advantages of high usability and strong empowerment ^[7]^.

**Table 8.**
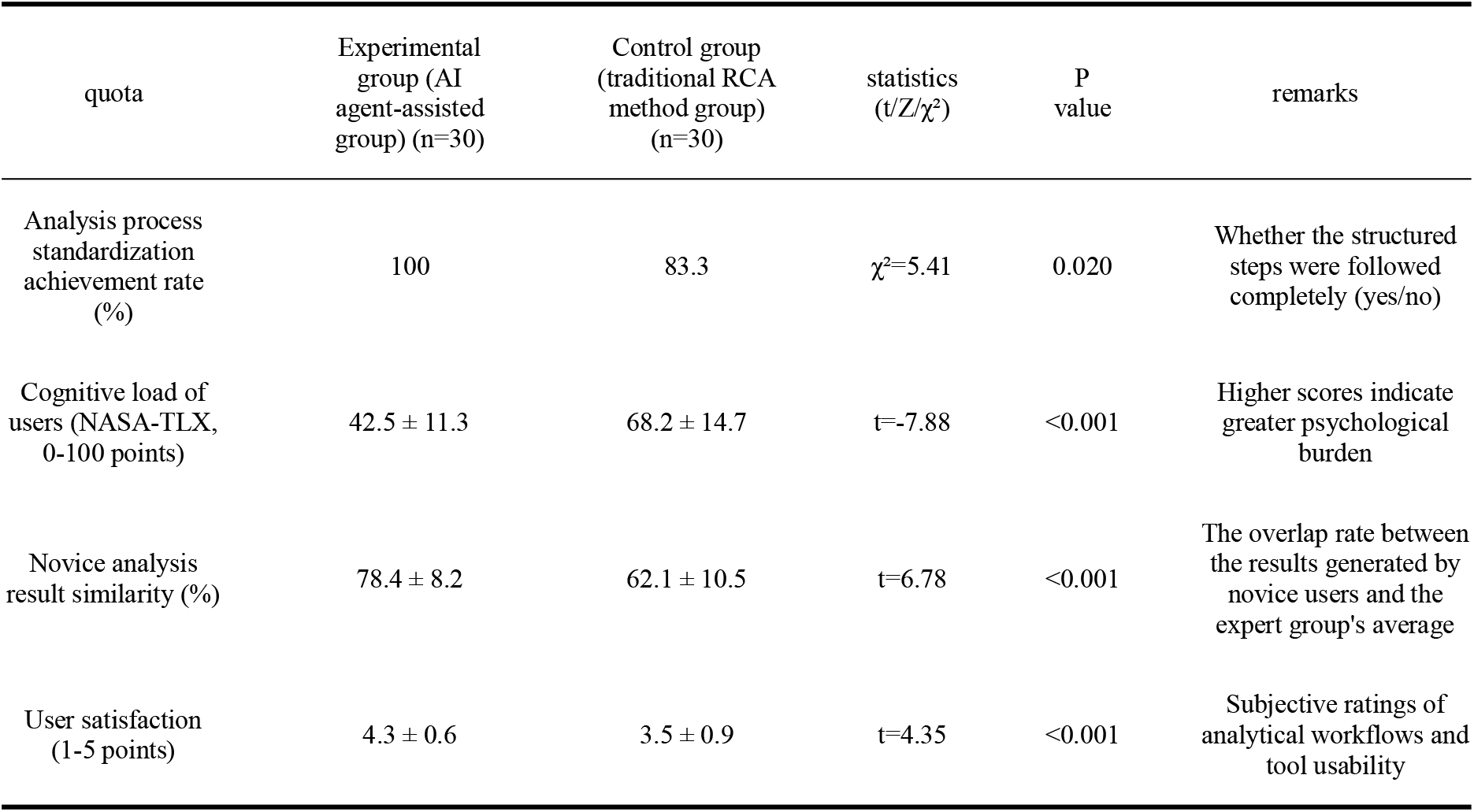
Process and User Experience Metrics Comparison.

| quota | Experimental<br>group (AI<br>agent-assisted<br>group) (n=30) | Control group<br>(traditional RCA<br>method group)<br>(n=30) | statistics<br>(t/Z/$\chi^2$) | P<br>value | remarks |
| --- | --- | --- | --- | --- | --- |
| Analysis process<br>standardization<br>achievement rate<br>(%) | 100 | 83.3 | $\chi^2=5.41$ | 0.020 | Whether the structured<br>steps were followed<br>completely (yes/no) |
| Cognitive load of<br>users (NASA-TLX,<br>0-100 points) | 42.5 ± 11.3 | 68.2 ± 14.7 | t=-7.88 | <0.001 | Higher scores indicate<br>greater psychological<br>burden |
| Novice analysis<br>result similarity (%) | 78.4 ± 8.2 | 62.1 ± 10.5 | t=6.78 | <0.001 | The overlap rate between<br>the results generated by<br>novice users and the<br>expert group's average |
| User satisfaction<br>(1-5 points) | 4.3 ± 0.6 | 3.5 ± 0.9 | t=4.35 | <0.001 | Subjective ratings of<br>analytical workflows and<br>tool usability |

### 2.5 Comparison of cost-effectiveness indicators between the two groups

Table 9 demonstrates that AI-assisted methods significantly outperform traditional approaches in cost-effectiveness metrics, particularly in single-session analysis time and training costs, achieving approximately 43% time savings and 50%-75% reduction in training duration. While requiring initial tool investment, their long-term benefits are substantial, as they enable continuous analysis quality optimization through knowledge accumulation and model iteration, ultimately forming reusable organizational intelligence assets. In contrast, traditional methods, despite lower upfront costs, rely on individual experience accumulation and struggle to achieve scalable improvements. The AI-assisted model enhances efficiency, lowers learning barriers, and strengthens organizations’ capacity for sustainable improvement, demonstrating superior overall cost-effectiveness. This makes it particularly suitable for scenarios with high standardization requirements and frequent analytical needs.

**Table 9.**
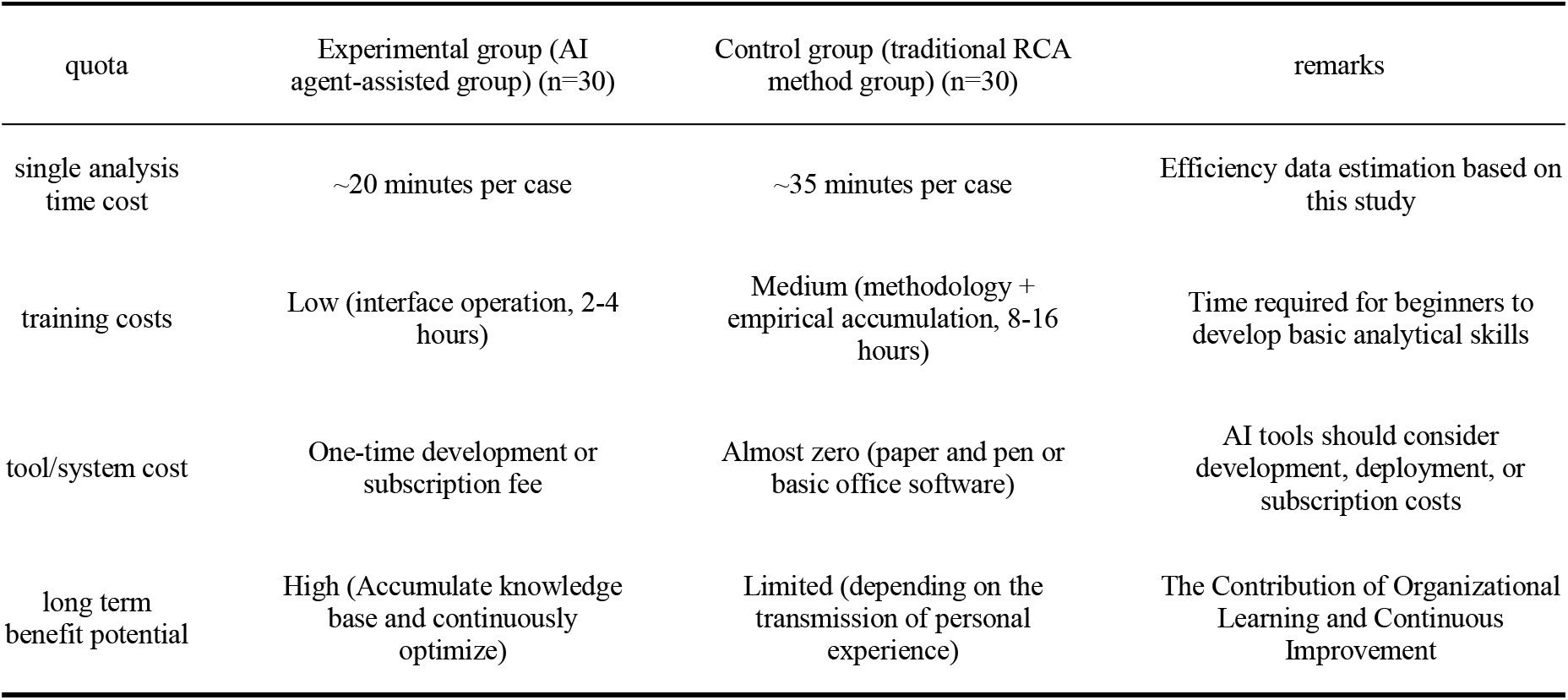
cost-benefit preliminary comparison.

| quota | Experimental group (AI agent-assisted group) (n=30) | Control group (traditional RCA method group) (n=30) | remarks |
| --- | --- | --- | --- |
| single analysis time cost | ~20 minutes per case | ~35 minutes per case | Efficiency data estimation based on this study |
| training costs | Low (interface operation, 2-4 hours) | Medium (methodology + empirical accumulation, 8-16 hours) | Time required for beginners to develop basic analytical skills |
| tool/system cost | One-time development or subscription fee | Almost zero (paper and pen or basic office software) | AI tools should consider development, deployment, or subscription costs |
| long term benefit potential | High (Accumulate knowledge base and continuously optimize) | Limited (depending on the transmission of personal experience) | The Contribution of Organizational Learning and Continuous Improvement |

## 3 discussion

The application of AI agents in complex decision-making scenarios is progressively overcoming the limitations of traditional methods. Its advantages are not only reflected in efficiency improvement and cost reduction, but also in promoting the standardization of analytical processes and the systematic accumulation of knowledge. The results of this study show that the AI-assisted group achieved a 100% compliance rate in analytical standardization, significantly higher than the 83.3% of the traditional group, indicating that AI-guided structured workflows effectively reduced human oversight and operational arbitrariness. Notably, novice users under AI support produced results with 78.4% similarity to expert panels, far exceeding the 62.1% in traditional modes, demonstrating that AI not only narrowed capability gaps but also enhanced result consistency and reliability. This phenomenon validates AI’s role as a “cognitive scaffolding” —externalizing expert logic to lower task execution thresholds. In contrast, traditional methods heavily rely on individual experience accumulation, with long training cycles and unstable outcomes, creating significant bottlenecks in organizational capacity building. Although current AI solutions involve certain initial investments, they exhibit significant cost-benefit advantages in long-term applications. Single analysis time savings of nearly 40% allow resources to be redirected toward higher-order cognitive tasks, while the system’s continuous learning and iteration capabilities enable the accumulation of organizational knowledge assets. Improved user satisfaction also reflects the positive impact of human-machine collaboration on work experience, with significantly reduced psychological burdens further indicating that AI alleviates procedural cognitive loads. While traditional methods offer the advantage of zero tool costs, their hidden costs—such as time-consuming training and inconsistent results—cannot be ignored in the context of growing demands for scalability and standardization. Our findings further support the view that technology assistance can narrow the “practical gap” between novices and experts ^[6]^. Future research should focus on balancing AI interpretability with user trust. Meanwhile, this study observed the adaptive potential of AI systems in cross-domain applications, where their modular architecture enables rapid deployment across decision-making scenarios, significantly enhancing organizational responsiveness. In contrast, traditional knowledge transfer heavily relies on personnel mobility and repetitive training, resulting in lower efficiency and potential distortion. Although some respondents cautiously questioned the absolute authority of AI recommendations, data shows that human-machine collaboration still yields better comprehensive judgment quality than single-subject independent decision-making. This suggests future development should focus on building transparent, traceable AI reasoning pathways to enhance user understanding and critical adoption capabilities. In the long term, AI is not just a tool replacement but a driving force for upgrading organizational cognitive paradigms. By making tacit knowledge explicit, AI is gradually reshaping the fundamental logic of knowledge management.

Traditional methods face limitations in achieving stable knowledge reuse due to individual memory biases and experiential gaps. AI systems, however, effectively mitigate execution drift caused by human factors through standardized processes, ensuring replicability at critical decision points. Yet in domains requiring comprehensive consideration of human factors and contextual complexity, traditional approaches retain their flexibility advantage, particularly evident in handling unstructured problems. While AI can provide data-driven recommendations, it still cannot fully replace human intuition in value balancing, ethical judgment, and dynamic relationship management.

The ideal future model should be a complementary human-machine collaboration, where AI assists in analysis to propose improvements, while human experts conduct in-depth evaluations, value calibration, and ethical considerations. This collaborative mechanism preserves the humanistic dimension of decision-making while enhancing overall efficiency and consistency. Human capabilities in complex scenarios—such as value judgments, ethical deliberations, and cross-domain associations—complement AI’s strengths in efficient computation and pattern recognition. This synergy not only optimizes resource allocation efficiency but also drives organizations to transition from reactive responses to proactive predictions. As technology matures, AI will progressively take on more foundational analytical tasks, freeing up human resources to focus on strategic innovation and emotional engagement. Ultimately, this human-machine symbiotic intelligent ecosystem will reshape organizational paradigms, achieving comprehensive upgrades across the entire knowledge creation, transmission, and application chain.

## Data Availability

All data produced in the present work are contained in the manuscript

## ACKNOWLEDGEMENTS

The authors thank the staff of the Central Sterile Supply Department at Sichuan Provincial People’s Hospital for their support in data collection and case validation. We also acknowledge the technical support provided by the Doubao AI platform team.

## Data Availability Statement

The data that support the findings of this study are available from the corresponding author upon reasonable request. The data are not publicly available due to privacy or ethical restrictions (anonymized quality management records from a single hospital).

## Author Contributions

Min Yi: conceptualization, methodology, formal analysis, writing – original draft.

Xinghua Zhang: software, validation, writing – review & editing.

Dan Zhao: data curation, investigation, formal analysis, writing – review & editing.

Qing Zhao: formal analysis, writing – review & editing, critical revision.

## References

[1] Lin M C, Yang C H, Chen Y J. An action research study of quality improvement in instrument packaging procedures for the central sterile supply department[J]. Scientific Reports, 2024, 14(1):2345.

[2] 中国信息通信研究院. 智能体技术白皮书 [R]. 2025.

[3] 江华, 顾宇峰, 蔡霞, 等. 基于人工智能的消毒供应中心灭菌质量管理系统的研发与应用[J]. 中国医学装备, 2020, 17(2):99–101.

[4] 郝艳丽, 代红红, 孙远, 等. 人工智能系统在外来医疗器械交接中应用效果的多中心研究[J]. 中华医院感染学杂志, 2023, 33(2):291–294.

[5] Wang Y, Li X, Zhang H. A FOCUS-PDCA quality improvement model for reducing the distribution defect rate of sterile packages[J]. Scientific Reports, 2023, 13(1):42295.

[6] Sixin J, Liangying Y, Yanhua C, et al. Optimizing Sterilization Packaging through Root Cause Analysis: An Exploration into Sealing Defects of Paper-Plastic Pouches[J]. Medical Science Monitor, 2023, 29:e940342.

[7] Raj M C, Osmani M A, Kuma T K. Quality improvement initiative in CSSD at a maternity care hospital using the PDCA cycle to support Kaizen (continuous improvement)[J]. International Journal of Applied Research, 2021, 7(8):1–5.

[8] 李睿男. 以缺陷管理改进模式为指导的质量管理在提升消毒供应室医疗器械清洗消毒合格率的影响[J]. 药物与人, 2025, 19(9):1–5.

[9] 关宁笑, 崔倩, 彭怡馨, 等. 中国医院消毒供应中心质量管理现状的 Meta 分析[J]. 中华医院感染学杂志, 2023, 33(1):1–8.

[10] 国家卫生和计划生育委员会.医院消毒供应中心第 1 部分:管理规范:WS310.1 一 2016[S]. 北京:中国标准出版社, 2017.

[11] 国家卫生和计划生育委员会.医院消毒供应中心第 2 部分:清洗消毒及灭菌技术操作规范:WS310.2-2016[S]. 北京:中国标准出版社, 2017.

[12] 国家卫生和计划生育委员会.医院消毒供应中心第 3 部分:清洗消毒及灭菌效果监测标准:WS310.3 一2016[S]. 北京:中国标准出版社, 2017.

[13] Hart G S. Nasa-Task Load Index (NASA-TLX); 20 Years Later[J]. Proceedings of the Human Factors and Ergonomics Society Annual Meeting, 2006, 50(9):904–908. DOI:10.1177/154193120605000909.

[14] Astina B A, Adrian Q J, Pasha D. User Interface Design of Mykonter Mobile Application Using User Centered Design Method on Konter Sam Cell[J]. Electronic Integrated Computer Algorithm Journal, 2024, 1(2):57-62. DOI:10.62123/enigma.v1i2.16.

